# A Hybrid Catheter Localisation Framework in Echocardiography Based on Electromagnetic Tracking and Deep Learning Segmentation

**DOI:** 10.1101/2020.12.22.20248705

**Authors:** Fei Jia, Shu Wang, V. T. Pham

## Abstract

Interventional cardiology procedure is an important type of minimally invasive surgery that deals with the catheter-based treatment of cardiovascular diseases, such as coronary artery diseases, strokes, peripheral arterial diseases and aortic diseases. Ultrasound imaging, also called echocardiography, is a typical imaging tool that monitors catheter puncturing. Localising a medical device accurately during cardiac interventions can help improve the procedure’s safety and reliability under ultrasound imaging. However, external device tracking and image-based tracking methods can only provide a partial solution. Thus, we proposed a hybrid framework, with the combination of both methods to localise the catheter tip target in an automatic way. The external device used was an electromagnetic tracking system from North Digital Inc (NDI) and the ultrasound image analysis was based on UNet, a deep learning network for semantic segmentation. From the external method, the tip’s location was determined precisely, and the deep learning platform segmented the exact catheter tip automatically.

## 1. Introduction

A cardiac interventional procedure, also known as an interventional cardiology procedure, is an important type of minimally invasive surgery that deals with catheter-based treatment of cardiovascular diseases, such as coronary artery diseases, strokes, peripheral arterial diseases and aortic diseases [1]. Generally, it can be classified into the following categories: cardiac catheterization, percutaneous coronary intervention, stents, embolic protection, percutaneous valve repair, balloon valvuloplasty and atherectomy. Catheter is the medical device used in most cardiac interventions that can be inserted into the body, which functionally allows for drainage, administration of fluids or gases, ablation and other tasks [2]. There are various types of catheters aiming at different medical applications, for example, the ablation catheter is specifically used for tissue ablation with the generated heat on the electrodes, the pacemaker catheter is to help heart pump, a central venous catheter is a conduit to give drugs positioned either in a vein near the heart or inside the atrium.

Image guidance during cardiac intervention is a key concept to guarantee patient safety while the direct line of sight is inhibited. X-ray imaging, traditionally, dominates the guidance during cardiovascular interventional procedures, but it provides limited views when the interventions involve the myocardium, pericardium and cardiac valves. Therefore, cardiac ultrasound (echocardiography) was introduced to navigate these challenges. Compared to cardiac X-ray imaging, echocardiography is especially useful for transcatheter puncture, percutaneous mitral valve procedures and left atrial appendage closure. Echocardiography fulfils the requirements by providing a real-time imaging solution, with both device and cardiac inner structure demonstration simultaneously [3]. There are three types of echocardiography that can be used during intervention: Transthoracic echocardiography (TTE), intracardiac echocardiography (ICE) and transoesophageal echocardiography (TEE/TOE). TTE is widely available and portable. It is a non-invasive imaging procedure [4]. However, it possesses limited ability to visualise the back of the heart and is difficult to use during interventional procedures. ICE has also demonstrated great potential for in-vivo medical device monitoring, where a thin probe is inserted inside a patient, but this phased array probe is expensive and can only be used once. Additionally, ICE offers no standard views [5]. As a trade-off between the image quality and imaging cost obtained through an echocardiography, TOE imaging is commonly chosen during the catheter-based intervention. Prior to imaging, the patient lies in the left lateral decubitus position, and swallows the probe following the instruction during probe insertion. Mild to moderate sedation is induced in the patient to ease discomfort and to decrease the gag reflex by providing medications, such as midazolam. This makes the ultrasound probe pass easily into the oesophagus.

Currently, 2-D multiplane imaging is the most widely used mode of TOE, providing 20 standard trans-oesophageal echocardiographic views that can facilitate and provide consistency in training, reporting, archiving and quality assurance (as published by the American Society of Echocardiography (ASE) and the Society of Cardiovascular Anaesthesiologists (SCA)) [6]. In clinical practice, before localising and tracking the device from echocardiography, a specific standard view should be determined first. For instance, to view the general four chambers, the probe is positioned at the mid-oesophagus with a zero degree rotation. It is then placed at the same position with a 40 degrees rotation. The aortic valve short-axis view can be obtained when the probe goes deeper into the stomach. The right ventricle and left ventricle views can be obtained at the same time from the transgastric apical short-axis view.

To ensure that the catheter tip is accurately localised during a safe interventional procedure when obtaining TOE imaging views, a reliable tracking solution is required. Currently, solutions, in general, can be categorized into two classes: the external tracking system and image-based method. The external tracking system needs to utilise an extra device to determine the catheter tip location; for example, the Bard Access product that employs the tip confirmation system (TCS) displays different electrocardiogram (ECG) signals, corresponding to different catheter locations [7]. However, surgeons require additional time and knowledge to analyse the external device and, occasionally, these external devices are largely affected by clinical environments. In comparison with external tracking methods, an image-based method is more distinct and easier to apply. Consequently, in recent years, the image-based method has attracted a lot of research attention. Previously, many image-based catheter tracking algorithms were performed on X-ray datasets instead of echocardiography datasets because X-ray images, electrodes or catheter tips possess distinct characteristic features that can be used for tracking and detection. At the same time, these features were vague in an ultrasound, which led to difficulties in localisation using only the image-based method. The classic image-based methods could only be applied on a small number of images. The methods utilised hand-crafted features. On the contrary, with deep learning, image tasks of greater difficultly can be achieved by end-to-end convolutional neural networks (CNN) [8]. Recently, the use of deep learning has been increasing rapidly in the medical imaging field, including computer-aided diagnosis (CAD), radiomics and medical image analysis [9].

However, the previous external tracking and image-based methods were two distinct localisation solutions to determine the catheter tip and no combination of these two methods was proposed with previous research studies.

The echocardiography images are collected on a 3D-printed, tissue-mimicking cardiac phantom [10] obtained from several standard TOE views with the Philips IE33 ultrasound machine [11]. Prior to ultrasound imaging, the catheter tip is first localised by the NDI EM tracking systems [12] using a pivot calibration [13] with an error less than 0.1 mm. Following data collection, all echocardiography images are processed with the Python 3 platform, using the UNet [14] automatic segmentation kernel. This hybrid localisation network can provide a reliable reference for new sonographers and doctors during catherization.

## 2. Materials and Methods

### 2.1. Echocardiography Image Collection for Catheter Localisation Model Training

To obtain the catheter tip information, at the same time, instead of using real patient data, the cardiac interventional procedure was simulated on a 3-D printed Lay-Fomm 40 phantom, which can fully resemble an adult patient heart. During imaging, the Philips S5-1 broadband sector array probe was placed on top of the cardiac phantom, while the phantom was fixed at the bottom of a plastic water tank. Subsequently, several TOE standard views were acquired by probe manipulation, such as the upper aortic valve view (commonly chosen in a real patient case). During the simulation, the Philips IE33 ultrasound machine was set to full volume mode with each image acquisition lasting for five seconds. All the DICOM images were then exported to blank CDs and analysed with ITK-SNAP [15].

The corresponding ultrasound imaging results of the Shelley medical ablation catheter movements can be observed in Figure 1. While the horizontal line is the artefact, which will not be labelled in the following works. In Figure 2, we can observe from the echo image that the background contained both the ablation catheter and the cardiac structures. In the aortic valve, the visualisation of the catheter is not only affected by the valve structure but also affected by strong reverberations from the water tank. The low-image resolution also increases the difficulty to localise the catheter tip accurately.

**Figure 1.**
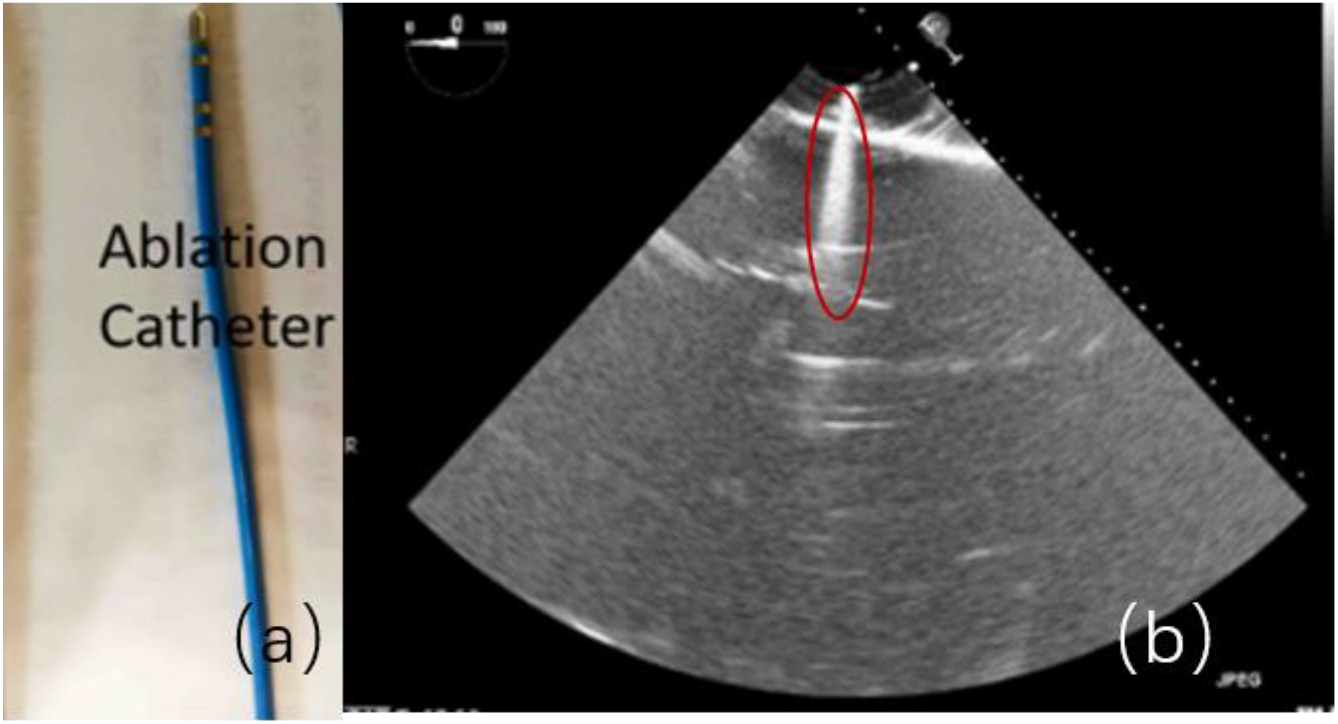
(a) Shelley Medical Ablation Catheter. (b) Corresponding 2-D Ultrasound Image of (a).

**Figure 2.**
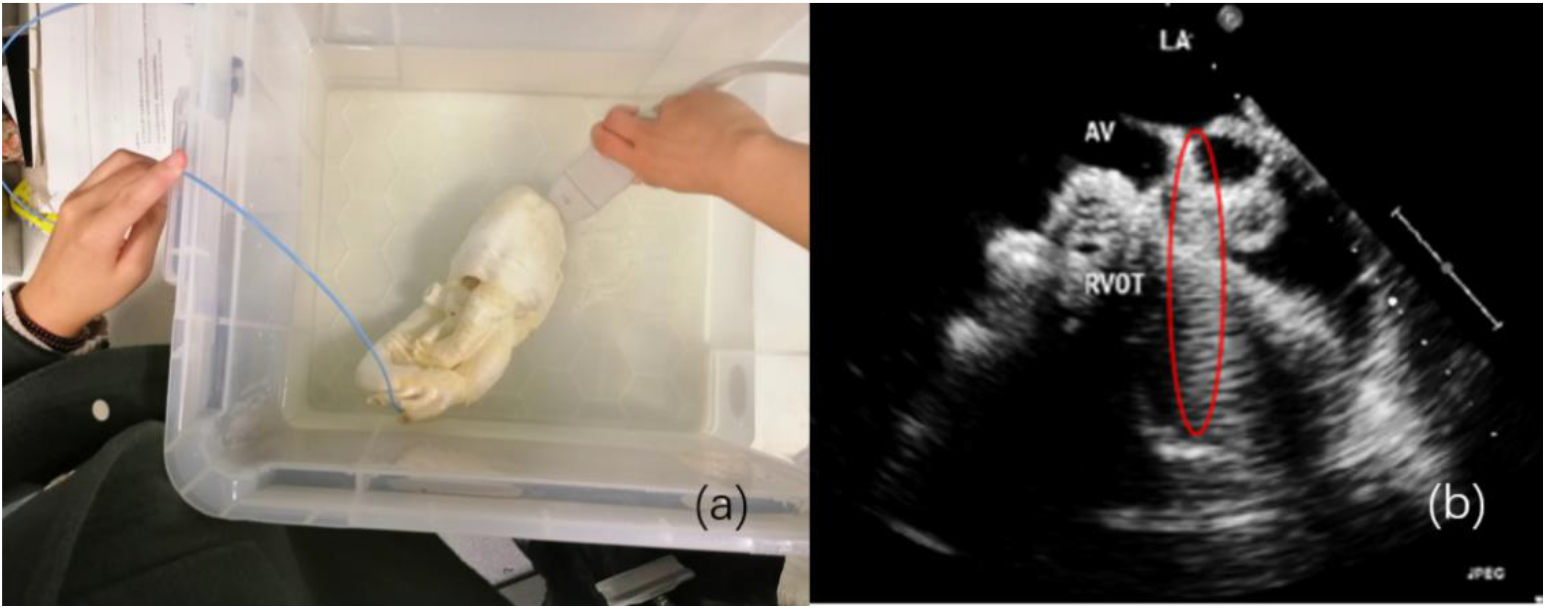
Catheter Tip Movement. (a) 2-D Echocardiography Image Acquisition on 3-D Printed Cardiac Phantom. (b) Corresponding Ultrasound Results of (a) Under Aortic Valve Short-Axis View

### 2.2. Catheter Tip Determination via External EM Device & Pivot Calibration

Before applying the deep learning network in image-based methods for automatic catheter segmentation, it was necessary to determine the exact location of the tracked catheter tip in the trained ultrasound dataset to provide the groundtruth. As illustrated in Figures 1 and 2, it is usually difficult to identify the catheter tip in the image by visual inspection alone. Therefore, mapping the physical location of the catheter tip to where it appears in the ultrasound imaging provided an alternative approach to obtain a reliable groundtruth.

In this section, the NDI Aurora EM tracking system [12] depicted in Figure 3 (a) was used to arrive at the catheter tip’s location physically because of its non-radiation and real-time 3-D tracking ability. To simplify the setup, a 6-degree of freedom (DOF) catheter-type EM sensor from NDI was used to represent the catheter, as it could generate a mapped point on a 2-D ultrasound image as indicated in Figure 3 (b). The Shelley medical ablation catheter could not be directly connected to the EM tracking device. It was tied to the sensor, so that the tracked tip location could be shared. The physical location of the tip was calculated through a pivot calibration experiment. The use of external EM tracking is to determine the length of catheter tip before training the lateral network, without EM tracking, the catheter tip cannot be defined during groundtruth labelling.

**Figure 3.**
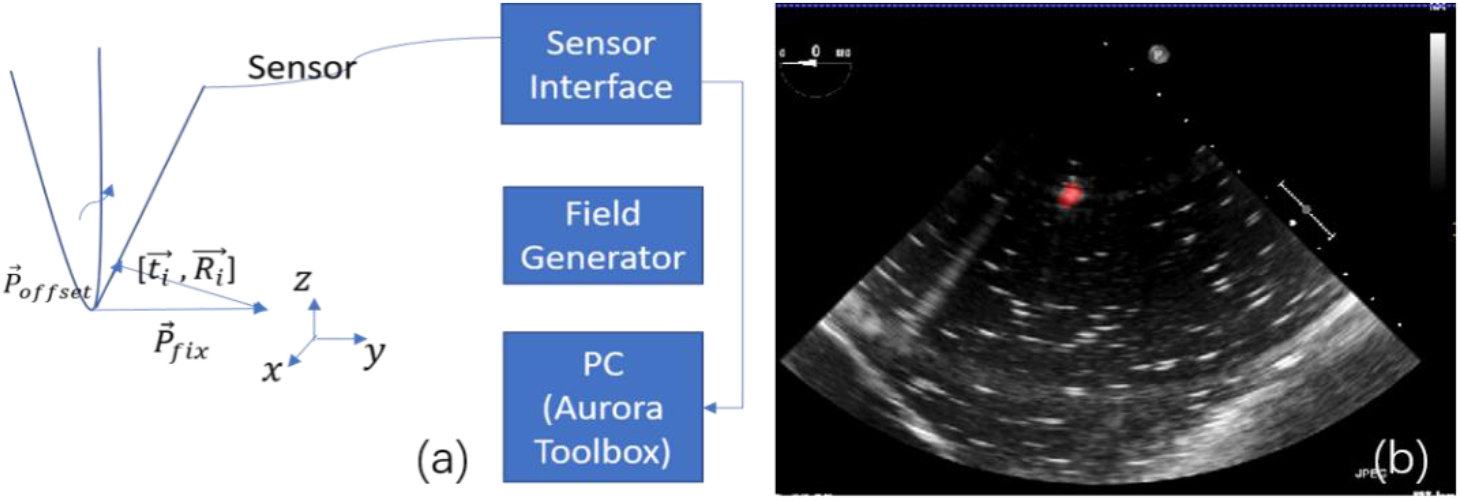
(a) Illustration of Pivot Calibration Using NDI Aurora EM Tracking System. (b) Mapped Location of Catheter Tip in 2D Echocardiography.

During tracking, all the data was recorded by the EM tracking device, saved as a text file and then processed further in MATLAB. Initially, the EM tracking device recorded the sensor’s 3-D location with the help of the tracking system coordinates. Through pivot calibration, the sensor was manually moved around a fixed pivot point near the EM field generator. The corresponding location matrix in the tracking system coordinates were transformed through formulas (1)–(5).

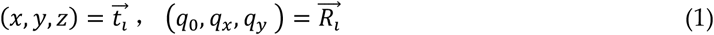

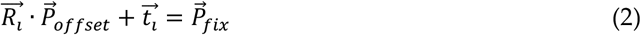

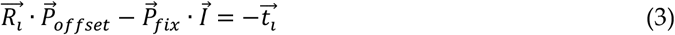

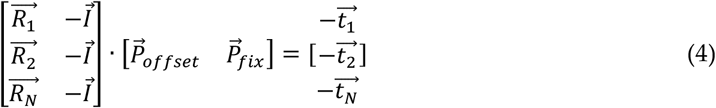

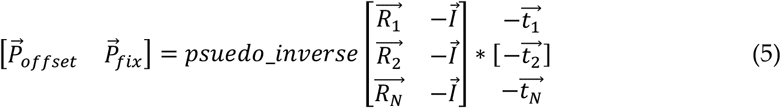

### 2.3. Automatic Catheter Segmentation in 2-D Echocardiography through Deep Learning

After determining the location of the ablation catheter tip physically, the state-of-the-art UNet was utilized to train the deep-learning-based automatic segmentation platform on the collected phantom echocardiography from different standard TOE views [16,17]. To make the trained model more robust, another 19 real patient TOE folders were mixed and tested at the same time.

Currently, the state-of-the-art semantic segmentation model being employed is UNet [18-23], which is depicted in Figure 4 [14]. This model can be used on smaller datasets, such as medical images for faster training, while the deep learning models have to be trained on larger datasets with more variations.

**Figure 4.**
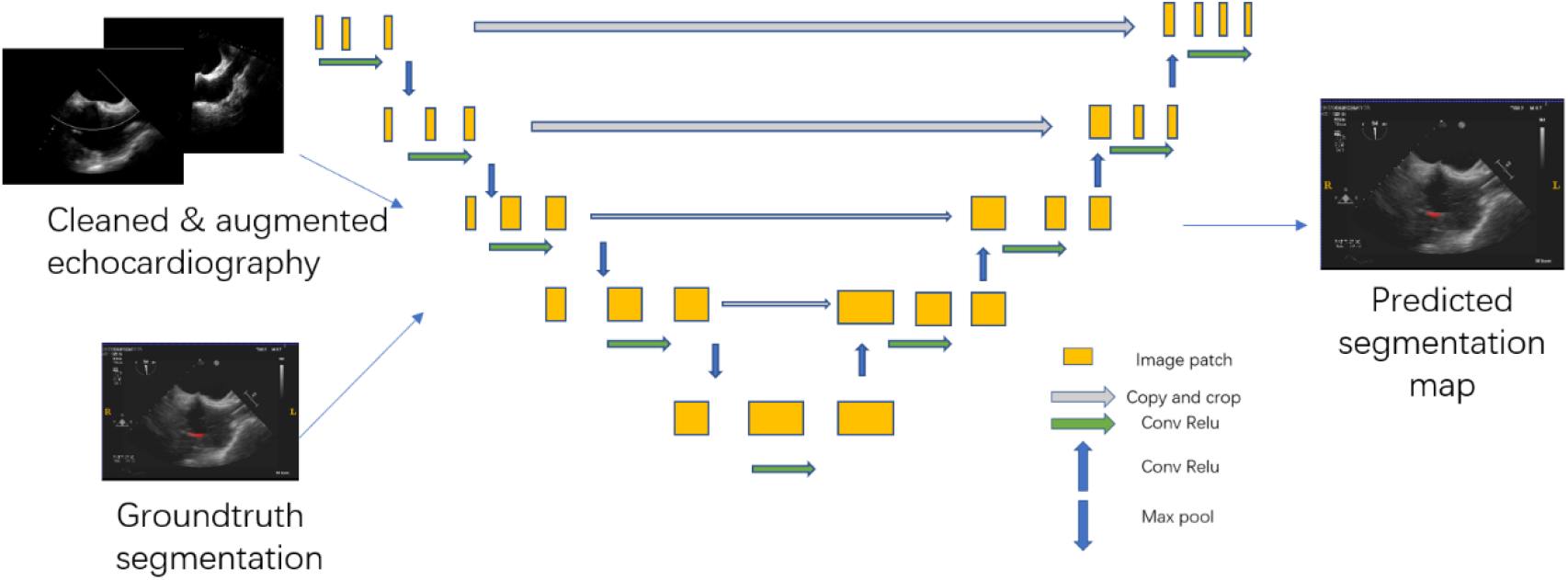
Diagram of Deep Learning Model Trained on 2-D Echocardiography Dataset.

The segmentation model was built on 2-D TOE images, collected from the Lay-Fomm 40 cardiac phantom, fabricated prior to obtaining both standard and non-standard views, with the ablation catheter moving from random places in the image. The image dataset contained 20 image volumes with 75 slices for each volume. The example provided in Figure 4 is a bicaval view with the catheter segmentation in the right atrium, and all of the groundtruth labels were obtained from the doctors’ manual segmentation (under the reference of both EM tracking results and visual inspections). The segmentation algorithm is written in Python 3. As illustrated from Figure 5, the model training procedure is given.

**Figure 5.**
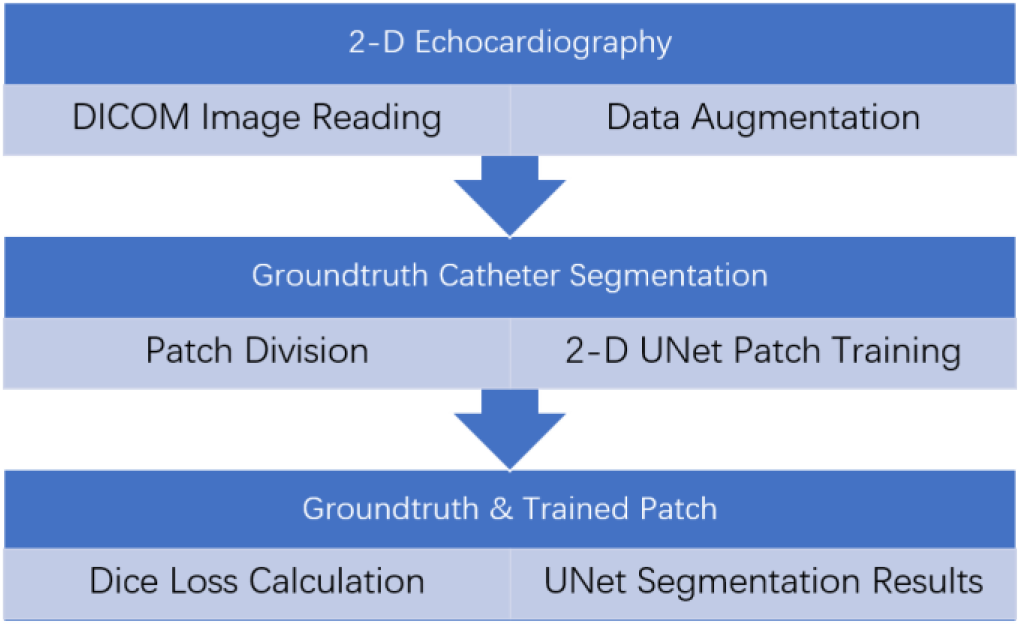
Flow chart of Echocardiography UNet Model Training Procedure.

The parameters used in the UNet segmentation model [24-32] were:

1. First ten volumes as the training dataset, second set of ten volumes as the validation dataset and testing on 14 random 2-D echocardiography volumes
2. Dropout at the last layer with the rate of 0.5
3. Augmenting the data offline to ten times as before
4. Use early stopping with patience of 6000 iterations
5. Positive and negative rate of 0.95 (when the image slice contained the segmentation target, we regarded it as a positive slice)
6. Patch size of 1,448,448
7. Batch size of four
8. Total number of iterations (batch size) were 30000
9. Iterative UNet depth of five
10. Loss function: Dice loss and Laplace smoothing for preventing overfitting
11. Activation function as ReLU

To better describe the performance by automatic segmentation when compared to groundtruth labelling, we introduced a Dice loss to evaluate how accurate the prediction would be (calculated through equation [37])

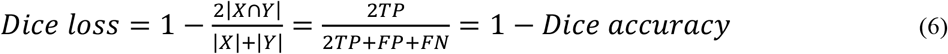

## 3. Results and Discussion

The physical localisation result of the targeted ablation catheter was 4.7362 ± 0.3523 mm, and the corresponding catheter’s groundtruth segmentation is indicated in Figure 6 (a). Figure 6 (b) indicates the corresponding prediction results of Figure 6 (a), trained by the deep learning platform. The original 2-D echocardiography of Figure 6 is the simulated result collected on the 3-D printed cardiac phantom. With the proposed hybrid framework, the accuracy of the catheter tip’s groundtruth location can be guaranteed to 0.1mm. When compared to the traditional groundtruth by considering a doctor’s visual inspection alone, this new groundtruth is more reliable. During the catheter movement, the final trained model could still identify the dynamic target as indicated in Figure 7. Within one second, no catheter tip was missing in every single frame, but at the same time, as the current deep learning network has limited ability in recognizing moving target, in figure 7, some predicted segmentations are incomplete compared to the groundtruth. The corresponding Dice accuracy results are as follows:

|  |  |  |
| --- | --- | --- |
| 0.8138811737712837 (max), | 0.6771465049473349, | 0.8079201691686618, |
| 0.7211444100551068, | 0.7322477650063857, | 0.7824308664136881, |
| 0.8112021032810474, | 0.6286978766145106 (min), | 0.7643511925952297 |

**Figure 6.**
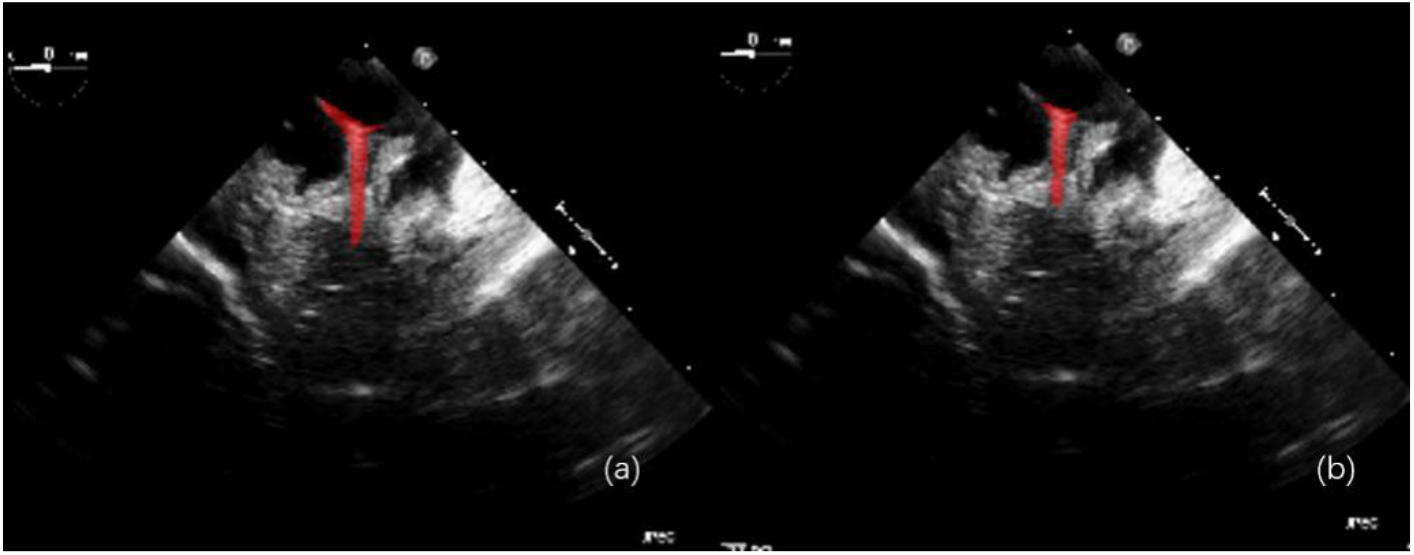
(a) Groundtruth segmentation of Shelley medical ablation catheter. (b) Deep learning platform predicted catheter segmentation on simulated 2-D echocardiography.

**Figure 7.**
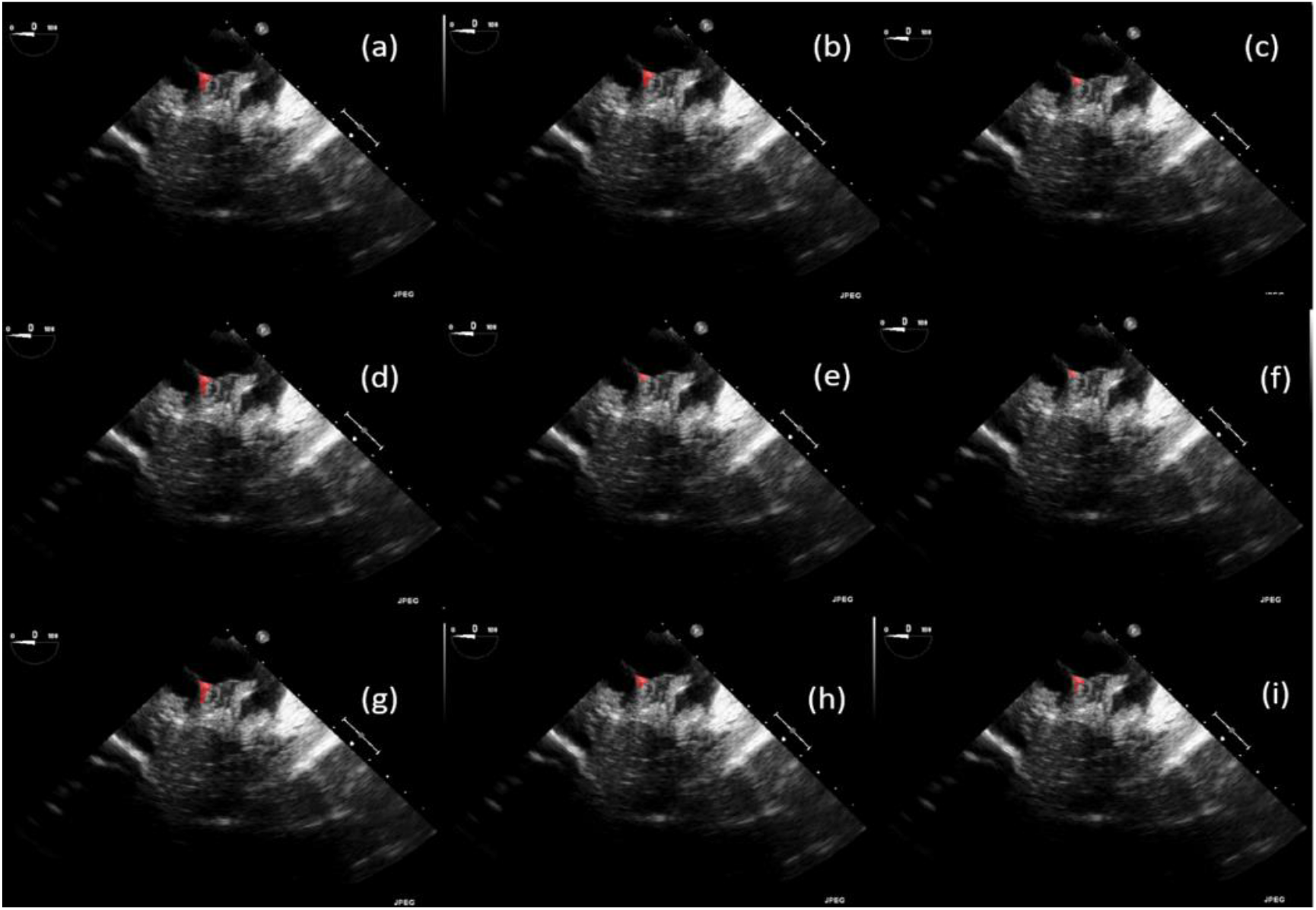
Catheter Tip Segmentation Results by Deep Learning Platform in 2-D Echocardiography Sequences (a) 0.1s (b) 0.2s (c) 0.3s (d) 0.4s (e) 0.5s (f) 0.6s (g) 0.7s (h) 0.8s (i) 1s

The training and testing of the Dice loss plots, indicated in Figure 8, are consistent with the aforementioned accuracy results. The Dice loss on training dataset was rather low. However, on the validation dataset, the Dice loss rose significantly (which indicated that the trained model was overfitting to a certain extent). Therefore, the accuracy obtained cannot compete with the performance on CT or MRI volumes. Except for the difficulty faced (due to the target being sparse or ambiguous), another challenge attributed to the limited variation of TOE images obtained, caused the overfitting of the model, which was unavoidable.

**Figure 8.**
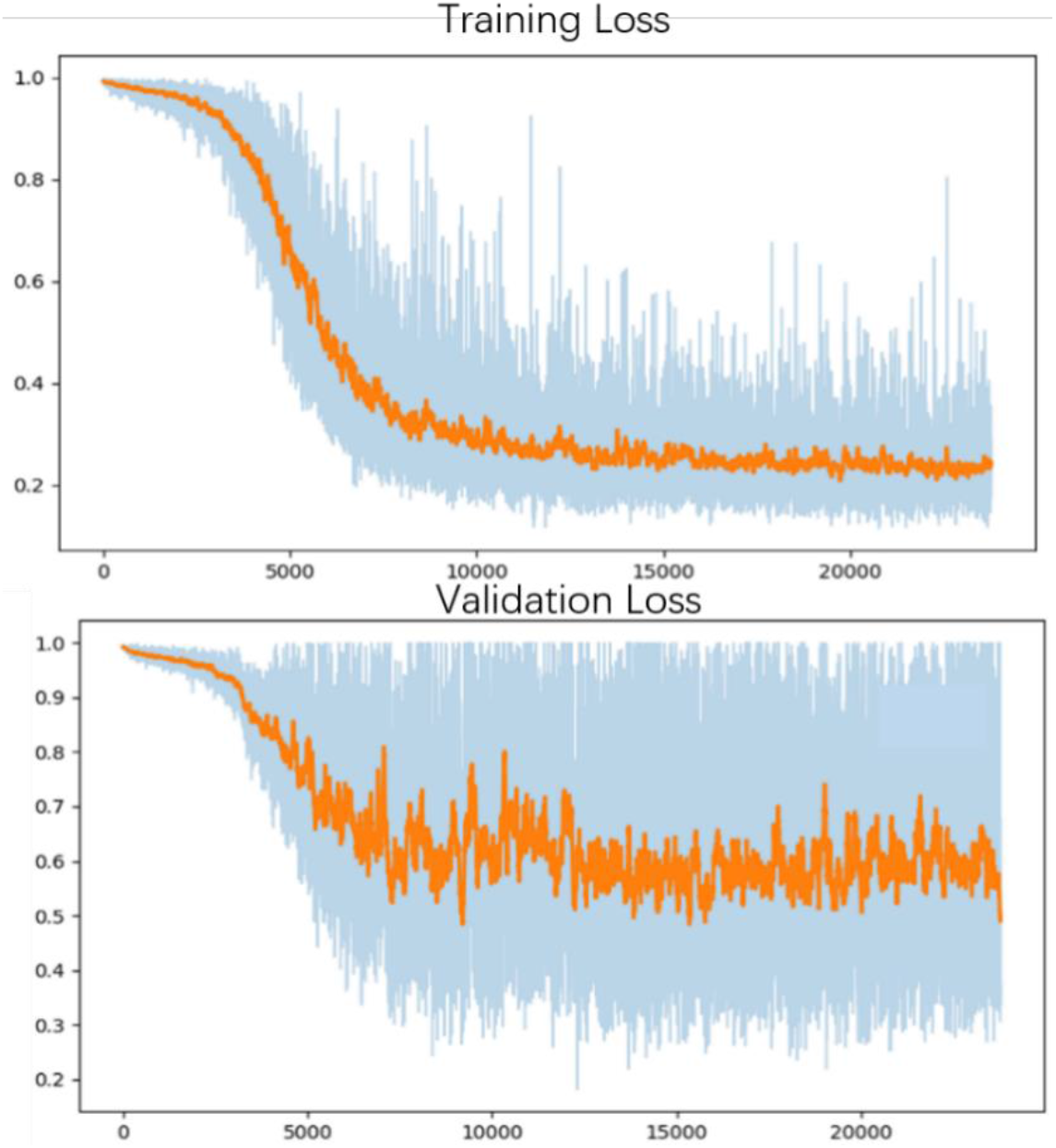
Training curve of the deep learning model’s training and validation loss.

To validate the trained model’s ability for generalisation, the model was also tested with real patient data obtained from several standard TOE views, such as the mid-oesophagus right ventricle (ME-RV) inflow view and the transgastric basal short-axis (TG basal SAX) view. From Figure 9, the predicted results proved the generalisation ability of the proposed model with correct catheter location and shape. As the target was too blurred, the shape may have varied from the groundtruth to a certain level.

**Figure 9.**
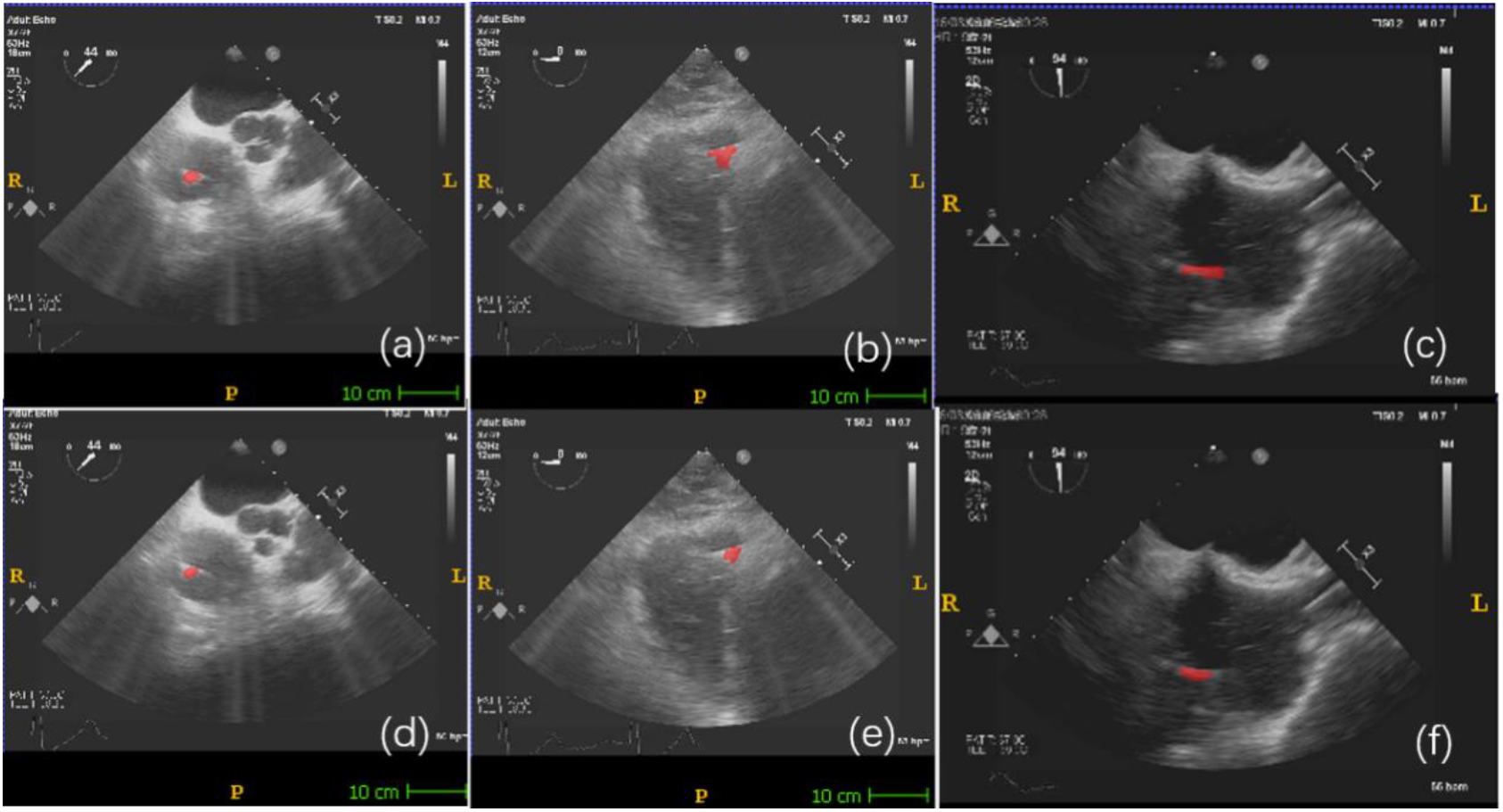
Validation of the model on real patient echocardiography (a)–(c): groundtruth segmentation from ME-RV inflow, TG Basal SAX and bicaval view. (d)–(f): deep learning predicted segmentation of (a)–(c)

## 4. Conclusions

EM tracking device is easily get affected by the clinical environment, and it cannot provide the visual information of the medical device; while the target in medical images cannot provide the numerical results. This new hybrid localisation framework combines the advantages of external EM tracking and deep-learning-based image methods, and successfully builds up the connection between the physical coordinate and the image coordinates, which offers a new solution for obtaining a more reliable groundtruth to train the automatic deep learning model. At the same time, 3-D printed phantoms also provide a new direction for collecting the original dataset to train the deep learning models based on our requirements.

Based on the simulated dataset and EM tracking tip determinations, the reliability of deep-learning-based models can be guaranteed. However, the model’s accuracy and stability need to be improved in the future. During the future improvement, the groundtruth labels have to be derived from the EM sensor, while all the possible standard views need to be classified too. Due to the dataset limitations, all the networks built thus far faced the overfitting problem, so an adequate fully automatic solution for cardiac intervention has not yet been achieved.

## Data Availability

The data in this statement can be accessed via the following links:
https://www.dropbox.com/s/75vc47oahi1tmkx/MDPI-images.zip?dl=0

## Data Availability

The dataset can be accessed upon request.

## Funding

This research received funding from the Chinese Scholarship Council and the Beijing Chuangxinhuizhi Co Ltd.

## Acknowledgments

We would like to express our gratitude for the financial support provided by the Chinese Scholarship Council and the Beijing Chuangxinhuizhi Co Ltd. We would also like to express our gratitude for the precious help provided by King’s College London.

## Conflicts of Interest

The authors declare no conflict of interest.

## Disclosure

An earlier version of this paper has been presented as preprint according to the following link: https://www.medrxiv.org/content/10.1101/2020.12.22.20248705.full.pdf [32].

## Authors’ Contributions

Fei Jia and Shu Wang designed the overall research scheme, programmed and debugged the algorithm, and wrote the paper. V. T. Pham made contribution in funding acquisition and proofreading.

## Notes

### Competing Interest Statement

The authors have declared no competing interest.

### Clinical Trial

The data used in this paper is obtained from phantom data, and the patient data is collected from the hospital ultrasound dataset, without new clinical trials.

### Author Declarations

The data in this paper doesn't involve any IRB approval, as we don't do any clinical trials on patient.

### Summary of Updates

New author Dr Vuongt Pham made contribution in funding acquisition and proofreading.

